# A comprehensive analysis of non-pharmaceutical interventions and vaccination on Ebolavirus disease outbreak: Stochastic modeling approach

**DOI:** 10.1101/2024.02.25.24302269

**Authors:** Youngsuk Ko, Jacob Lee, Yubin Seo, Eunok Jung

## Abstract

Ebolavirus disease (EVD) outbreaks have intermittently occurred since the first documented case in the 1970s. Due to its transmission characteristics, large outbreaks have not been observed outside Africa. However, within the continent, significant outbreaks have been attributed to factors such as endemic diseases with similar symptoms and inadequate medical infrastructure, which complicate timely diagnosis. In this study, we employed a stochastic modeling approach to analyze the spread of EVD during the early stages of an outbreak, with an emphasis on inherent risks. We developed a model that considers medical staff and unreported cases, and assessed the effect of non-pharmaceutical interventions (NPIs) using actual data. Our results indicate that the implementation of NPIs led to a decrease in the transmission rate and infectious period by 30% and 40% respectively, following the declaration of the outbreak. We also investigated the risks associated with delayed outbreak recognition. Our simulations suggest that, when accounting for NPIs and recognition delays, prompt detection could have resulted in a similar outbreak scale, with approximately 50% of the baseline NPIs effect. Finally, we discussed the potential effects of a vaccination strategy as a follow-up measure after the outbreak declaration. Our findings suggest that a vaccination strategy can reduce both the burden of NPIs and the scale of the outbreak.

**Author summary:** Our research employs a stochastic model to analyze the early-stage spread of Ebolavirus Disease. We incorporated factors such as medical staffs and unreported cases, and utilized real data to evaluate the impact of non-pharmaceutical interventions on disease transmission. Our findings indicate that rapid outbreak recognition could effectively control disease spread with reduced efforts. Furthermore, we explored the potential implementation of a vaccination strategy following an outbreak declaration. Our results suggest that such a strategy could mitigate both the scale of the outbreak and the necessity for additional interventions.

## Introduction

Ebolavirus Disease (EVD) was first identified in 1976 in Sudan and the Democratic Republic of Congo [1, 2]. The scale and impact of EVD outbreaks have evolved over time. The 2013-2016 West Africa epidemic was notably the most severe outbreak, resulting in over 11,000 deaths [3]. The most recent outbreak occurred in Uganda in 2022, with the first case identified on September 19, leading to an official outbreak declaration the following day. This outbreak lasted approximately four months, with 164 confirmed cases and 77 deaths [4]. The management and response to EVD outbreaks pose several challenges. For instance, cases may go unreported due to the initial symptoms being easily mistaken for other diseases, leading to transmission to medical staffs (MS) during the early stages of an outbreak [5, 6]. This challenge is particularly prevalent in remote areas and regions with limited medical facilities [7].

As of January 2024, two vaccines have been approved: Ervebo and Zabdeno/Mvabea [8]. Ervebo, a single-dose vaccine, is primarily used for emergency response and has demonstrated near 100% efficacy in preventing infection immediately after vaccination [9]. However, it is only effective against Zaire ebolavirus and has poor storage stability, requiring use within 4 hours at room temperature and temperatures below -60°C for long-term storage [10]. In contrast, Zabdeno/Mvabea, a two-dose vaccine administered to healthcare workers in advance, is speculated to have a relatively lesser preventive effect than Ervebo [10]. It requires an 8-week vaccination period for the two doses, making it unsuitable for immediate outbreak response, but it has better storage stability, remaining viable for up to a year at regular refrigerator temperatures [10].

The application of these vaccines during outbreaks offers further insights. During the 2013-2016 West Africa outbreak, ring vaccination commenced in April 2015, a period when the outbreak was subsiding. The vaccination was experimental, with a small number of approximately 3,000 individuals vaccinated compared to the overall scale of the outbreak, and the effectiveness of the vaccine was measured during the same period [11]. In the 2018-2020 Kivu epidemic, vaccination started a week after the outbreak was declared, and approximately 300,000 individuals were vaccinated in total [12]. However, despite rapid recognition and the application of both non-pharmaceutical interventions (NPIs) and vaccination, controlling the spread was challenging due to conflict, insecurity, and misinformation [13–15].

Utilizing mathematical modeling to study infectious diseases provides a systematic structure that is crucial for deciphering and forecasting disease transmission dynamics [16, 17]. A significant advantage of such modeling is its ability to provide quantitative insights. Rather than making decisions based on general observations, these models utilize detailed numerical data. This precise data assists policymakers in understanding the outcomes of potential interventions, including vaccination campaigns, travel restrictions, and the enforcement of social distancing measures [18–20]. Numerous studies have primarily focused on the mathematical modeling of transmission dynamics and control strategies related to EVD outbreaks. Previous research has investigated the initial transmission patterns during the 2014 West African EVD outbreak to quantify the disease’s transmissibility and the impact of NPIs [21, 22]. The potential risks associated with importing the pathogen into non-African countries and the inherent threats of large-scale outbreaks were examined [23, 24]. Several studies have utilized contact tracing strategies to assess the effectiveness of various containment and intervention approaches [25, 26].

Numerous studies have concentrated on vaccination strategies for EVD outbreaks. Masterson analyzed the required level of preventive vaccines based on the basic reproduction number within a population and concluded that the ideal vaccination coverage is unrealistic due to the high requirement [27]. Chowell used an individual-based model to evaluate the impact of vaccine strategies on outbreak control and found that ring-vaccination alone would not be effective in controlling the epidemic in situations where there is a delay in vaccination [28]. Wells conducted a spread analysis in Congo using a spatiotemporal model and observed that the vaccine program reduced the risk areas by up to 70.4% and decreased the risk level within those areas by up to 70.1% [29]. Lastly, Potluri found that if preventive vaccine strategies are applied to healthcare workers and the general population, the scale and mortality of EVD outbreaks can be significantly reduced, even considering only imperfect vaccine effects [30].

In this study, we investigated the early stages of the EVD outbreak, considering various key factors associated with potential risks. In modeling the EVD outbreak, we adopted a comprehensive approach by considering the roles of MS, unreported cases, and the lag between the emergence and detection of the initial case while assessing the effect of NPIs, including vaccination strategies. While some of the factors we incorporated for EVD have been investigated in previous research, our methodology is unique as it combines unreported cases, MS, NPIs, and vaccines into a single detailed model and distinctly measures the outcomes.

## Materials and methods

### Modeling of EVD outbreak

In the modeling of the EVD outbreak, we considered the following groups: susceptible (*S*), exposed (before symptom onset, *E*), infectious (post-symptom onset, *I*), hospitalized (*Q*), and recovered (*R*). We further divided the infectious group into *I*_1_ and *I*_2_ to differentiate between reported and unreported cases. We hypothesized that hospitalized patients were effectively isolated and could not transmit the disease. We incorporated MS by adding groups with the subscript *M*, and found that there were no unreported cases among the MS. Fig 1 outlines the entire progression of the disease. Solid arrows indicate infection events characterized by non-delayed reactions (Markovian processes). In contrast, dashed arrows denote disease progression and delayed reactions, which are non-Markovian processes.

**Fig 1.**
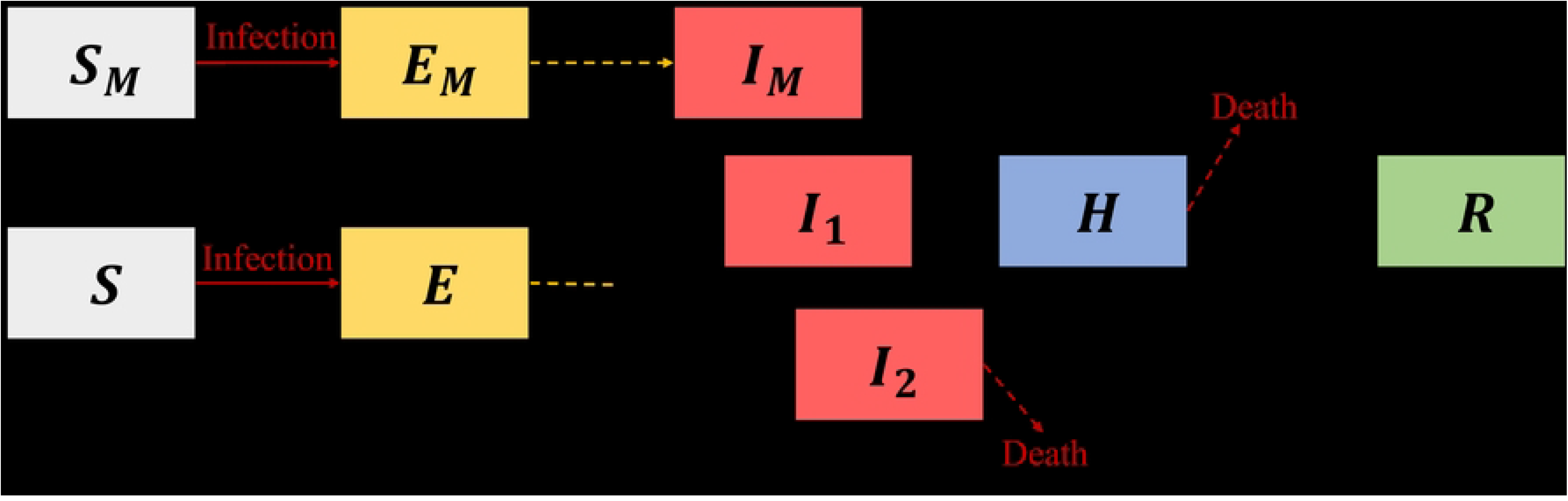
Flow diagram of the Ebolavirus disease transmission model. Medical staffs and unreported cases are considered. Solid-line arrows signify nondelayed reactions, whereas dashed-line arrows denote delayed reactions.

The non-delayed reactions in infection transmission are described as follows for both MS and non-MS:

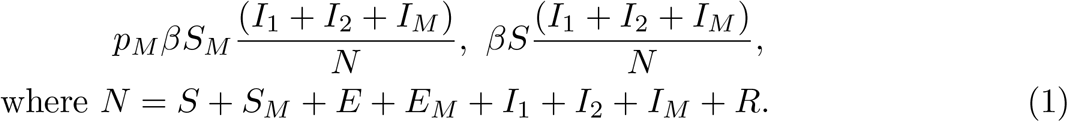

The parameter *β* is the transmission rate and *p*_*M*_ represents the heightened risk factor associated with MS. The parameter *p*_*M*_ is determined to be 254.55, ascertained from the case number ratio of non-MS to MS (50:14) and the population size ratio (1000:1.1) in Mubende province, where the study was conducted [31, 32]. These data indicated that MS poses a greater risk of infection by 254.55. We estimated the value of *β* without NPIs at 0.19, assuming that the basic reproductive number is 2.5 and the average infectious period is 5.79 days [33–35]. We set the case fatality rate *f* to 0.44 [36]. The subsequent subsection discusses the report rate, represented by *ρ*, which varies based on the outbreak detection.

We used modified Gillespie algorithm to simulate our model and ran simulation 10,000 runs per scenario [37]. Table 1 offers a comprehensive breakdown of the propensities associated with non-delayed events and the particulars of delayed events. We assumed a uniform distribution of delays, except for the incubation period, owing to a lack of data.

**Table 1.** Characteristics of the propensity of nondelayed events and details of delayed events.

| Event | Type | Description | Reference |
| --- | --- | --- | --- |
| Infection of MS | Nondelayed | Propensity:<br>$p_M \beta S_M \frac{I_1 + I_2 + I_M}{N}$ | [33–35] |
| Infection of non-MS | Nondelayed | Propensity:<br>$\beta S \frac{I_1 + I_2 + I_M}{N}$ | [31–35] |
| From exposure to onset | Delayed | Log-normal distribution,<br>Mean: 9, SD: 4.31 | [38] |
| Symptom onset to hospitalization (non-MS) | Delayed | Uniform distribution,<br>$5.79 \pm 3.30$ | [34, 35] |
| Symptom onset to hospitalization (MS) | Delayed | Uniform distribution,<br>$0.5 \pm 0.5$ | Assumed |
| From hospitalization to recovery | Delayed | Uniform distribution,<br>$20.38 \pm 7.58$ | [34, 35] |
| From hospitalization to death | Delayed | Uniform distribution,<br>$5.56 \pm 6.11^*$ | [34, 35] |

\*If the generated value is lesser than 0, then the value changes to 0. In real, this is the case when the patient dies before the hospitalization.

### Scenarios for model simulation

For a baseline scenario, we focused on the outbreak within the Mubende district, the epicenter of the 2022 Ugandan EVD outbreak. The simulation encompassed two stages, accounting for behavioral alterations and NPIs after the outbreak announcement: the phase before the declaration (*P* 1) and after the declaration (*P* 2). Fig 2 graphically describes and clarifies this phase division. A primary case refers to an individual introducing the infection into a population, whereas an index case denotes the first identified case [39]. The primary case can be the index case, but not necessarily.

**Fig 2.**
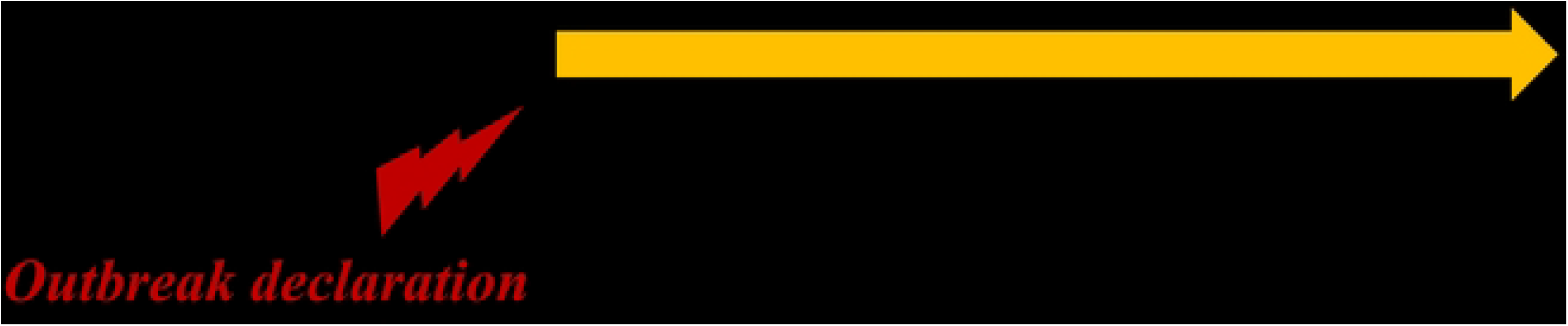
Division of phases considering outbreak declaration and setting for baseline model simulation scenario.

To set the effect of NPIs for the baseline scenario, we assumed that the transmission rate and duration from symptom onset to hospitalization decreased by 30% and 40%, respectively, upon outbreak declaration. Note that these coupled values (30% and 40%) are chosen to simulate real incidence and described in subsequent section. The criterion for this declaration was 19 days after the first death, which is within the reported group and consistent with the situation in Uganda. In 2022, it was ascertained in Mubende district that six deaths, later confirmed, had occurred before the official outbreak declaration [40, 41]. Investigations indicated the potential for 17 more probable deaths before this declaration [42]. Based on these data, the reporting rate in the pre-declaration phase was estimated to be 7/24 (29.17%). In the post-declaration phase, with 22 confirmed deaths and two probable deaths, the estimated rate was 22/24 (91.67%). The report rate also shifted (from *P* 1 to *P* 2) when the outbreak was declared. Furthermore, at the outbreak declaration, we assumed that previously unreported individuals are later reported based on the difference between the two reporting rates.

To consider comparable scenarios, we explored the effects of varying the thresholds for outbreak declaration, the intensity of NPIs on the spread of the disease, and vaccination. The key variations considered are as follows:

- Threshold for outbreak declaration: The delay from the first death to outbreak declaration varied from 1 to 38 days.
- Effect of NPIs: The effect of NPIs on the transmission rate and infectious period varied. This variation ranged from a 50% reduction (more stringent NPIs) to an increase of 50% (less severe NPIs) relative to the baseline setting.
- Vaccination: It is assumed that a person is immediately immune, i.e., hosts in state *S* transfer to *R*, once the vaccination is completed within a certain period (minimum 10 days, maximum 90 days) after the outbreak is declared. Proportion of vaccinated individuals is ranged from 0.1 to 0.3, whereas all of MS are vaccinated.

## Results

### Baseline scenario simulation

Fig 3 shows the baseline simulation results for cumulative confirmed cases. The gray curves depict the outcomes of each distinct simulation run, the dark curve signifies the mean, and the red boxes show the trends of confirmed cases in Mubende district. Because of inherent randomness, the timing of the outbreak declaration differs across runs; therefore, all simulation outcomes were synchronized based on the timing of the outbreak declaration. The actual number of confirmed cases in Mubende District was 66. The simulation mean value was 66.84, with a 95% credible interval (CrI) ranging from 0 to 226.

**Fig 3.**
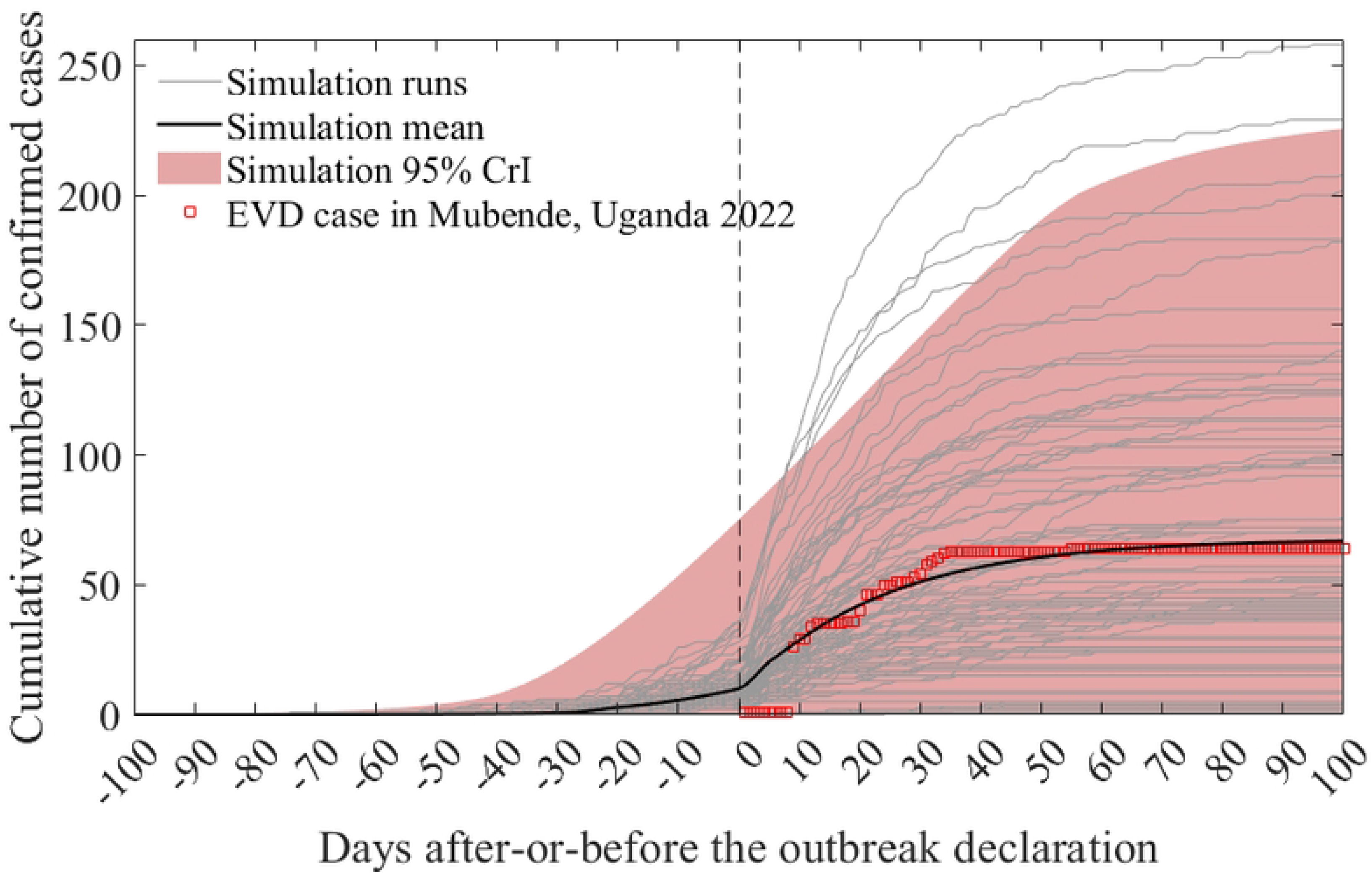
Cumulative confirmed cases from the baseline model simulation. The grey curves represent individual simulation runs, the dark curve denotes the simulation mean, and the red boxes display actual data from the Mubende district. Note that the vertical line, marking time 0, signifies the timing of the outbreak declaration in the simulation runs.

In the baseline scenario simulation, the transmission rate and duration from symptom onset to hospitalization (infection period) were reduced by 30% and 40%, respectively, following the outbreak declaration. Thus, the real-world effect of NPIs closely mirrors these levels. Nevertheless, the decline in the transmission rate might have been more pronounced, whereas the reduction in the duration from symptom onset to hospitalization might have been less significant or the inverse. Fig 4 shows the contour lines for pairs of values with an average closely aligned with the actual data, spanning a range of the effect of NPIs. Red asterisk indicates values for the baseline scenario (40% and 30% of reduction of infectious period and transmissibility, respectively). When comparing the X- and Y-axis intercepts, scenarios with no reduction in the infectious period but a 70% reduction in the transmission rate and those with no decrease in the transmission rate but a 53% reduction in the infectious period showed similar simulation results.

**Fig 4.**
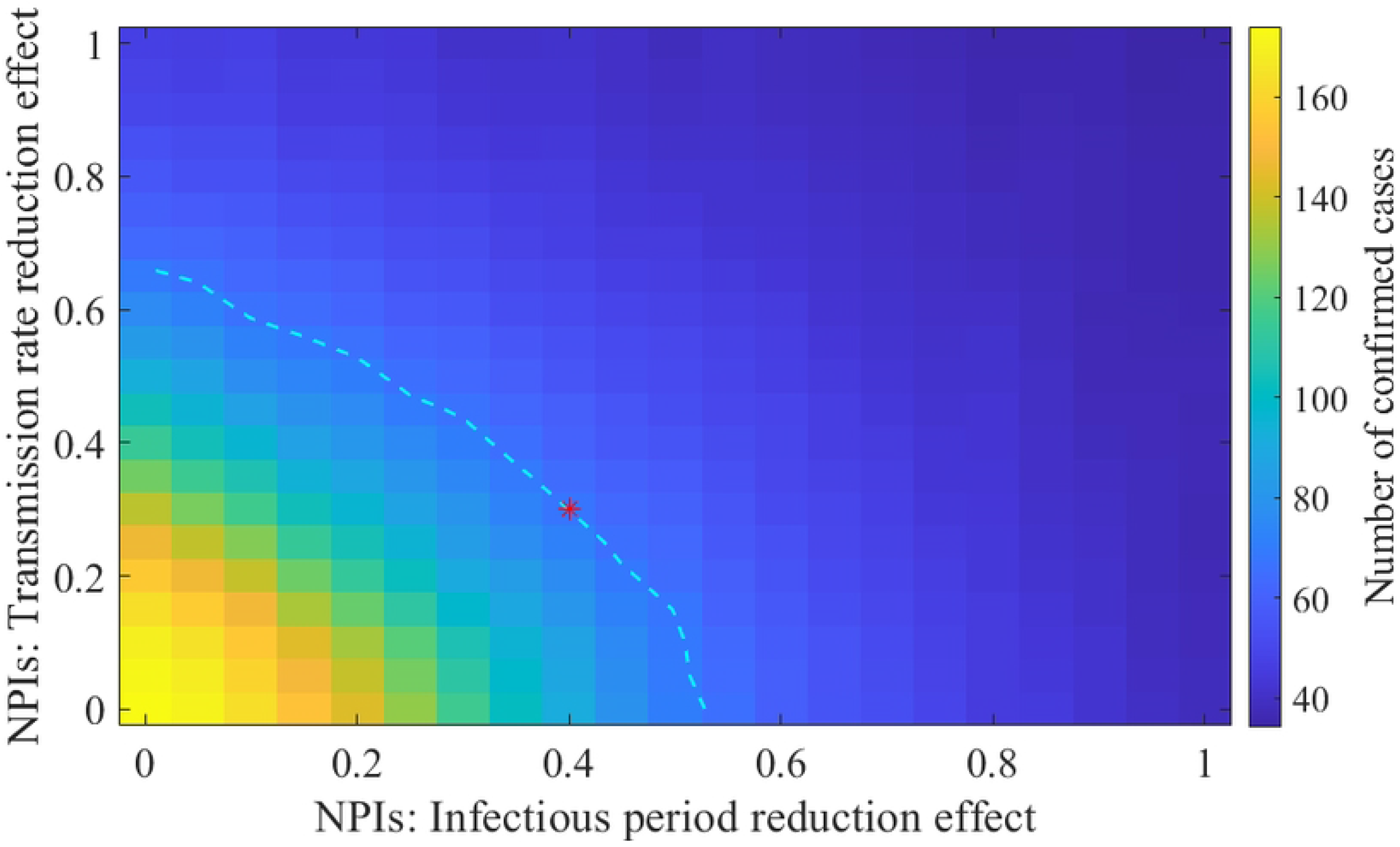
Effect of NPIs on the simulation results for confirmed cases. The dashed cyan curve represents the contour line with an average value equivalent to the baseline scenario simulation outcome.

Fig 5A presents the distribution of duration from primary case to outbreak declaration (*P* 1), which reveals a bimodal pattern. Outbreaks are typically declared within seven days, with another cluster emerging at approximately 50 days. This distinct pattern arises in some simulation scenarios where subsequent infections do not manifest, leading to the premature end of the outbreak. The probability of an outbreak concluding prematurely within one week was 19%. On average, excluding instances where the disease ended early, the primary case manifested approximately 50 days prior, with a 95% CrI ranging from 32 to 82 days. Fig 5B shows the distribution of *P* 2 duration and exhibits a monomodal distribution, with a mean of 64 days and a 95% CrI spanning from 19 to 152 days. In this study, we defined the duration of *P* 2 as the period from outbreak declaration to when there were no individuals in stages *E* or *I*.

**Fig 5.**
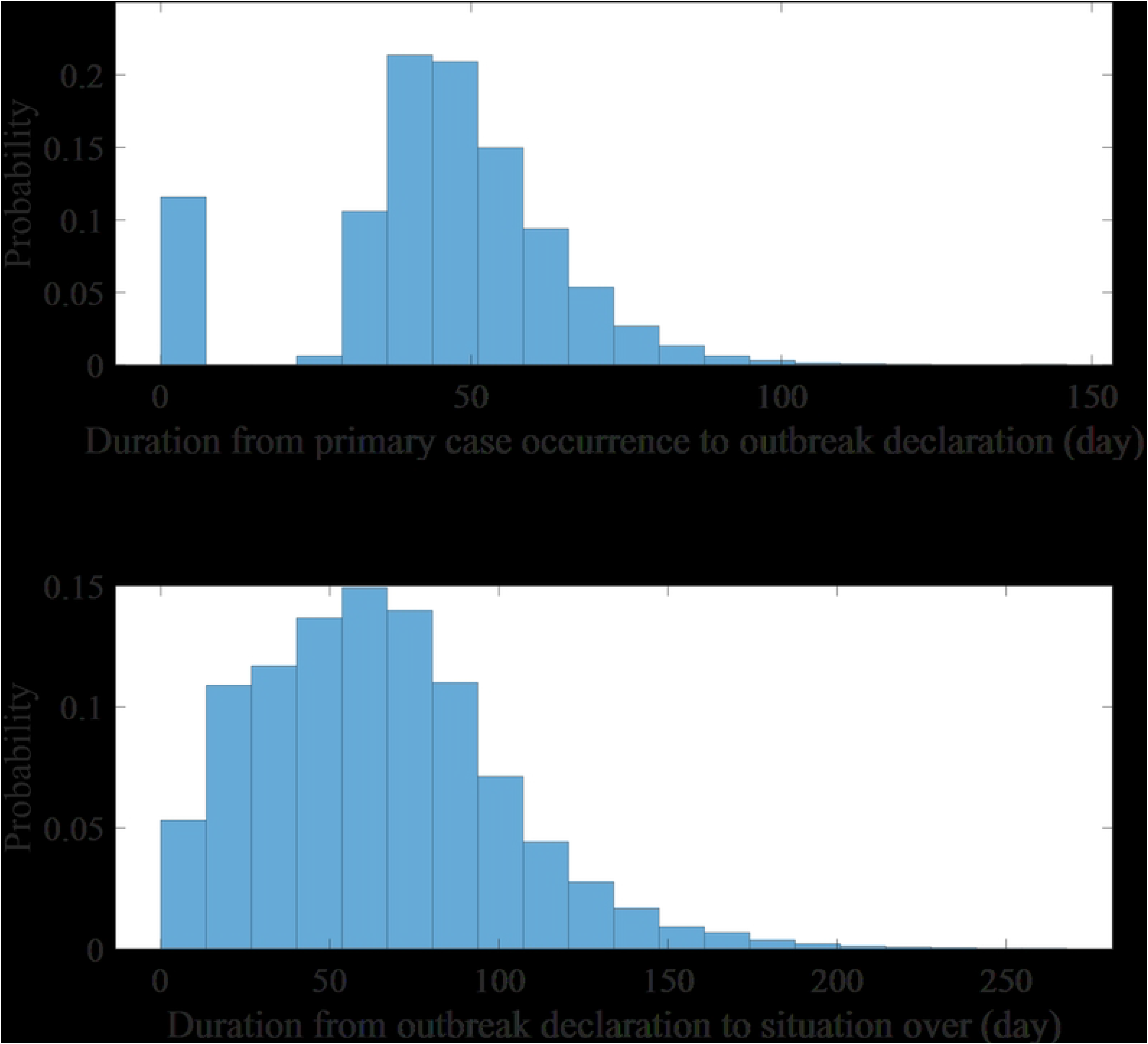
Histogram representing the durations of phases. Duration from the occurrence of the primary case to the outbreak declaration (A), and from the outbreak declaration to the end of outbreak (B).

Addressing how many individuals were infected when the outbreak declaration was officially acknowledged is essential for planning and responding. Fig 6 illustrates the distribution of prevalence by status at the outbreak declaration. Mean number (95% CrI) of *E, E*_*M*_, *I*_1_, *I*_2_, and *I*_*M*_ are 12.07, 3.04, 1.64, 7.34, and 0.16 ([0,45], [0,11], [0,7], [0,27], and [0,1]), respectively. When normalized by population size, the number of exposed MS is 27.62 per 1,000. This ratio is 229 times higher than the non-MS group, which registers at 0.12 per 1,000.

**Fig 6.**
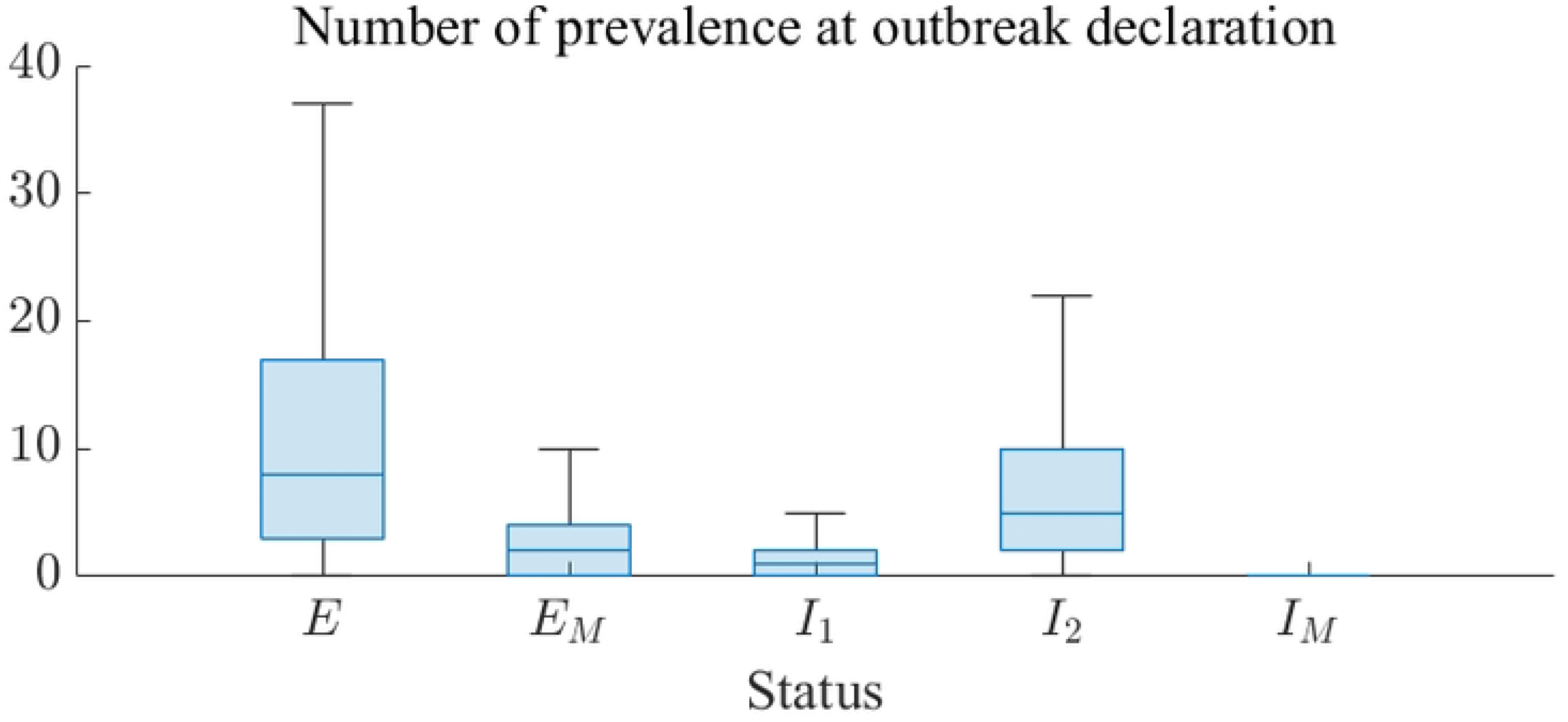
Distribution of prevalence by each status at the time of outbreak declaration.

### Scenarios considering NPIs and outbreak detection

We examined the distribution of confirmed case numbers across various settings, ranging from 1 to 38 days leading up to the outbreak declaration from the occurrence of the first death (or variations in NPIs levels ranging from -50% to +50% relative to the baseline). Fig 7A (Fig 7B) shows the delay range (NPIs levels) on the x-axis against the number of confirmed cases on the y-axis. As expected, with an increase in the delay, the number of infections also increased, exhibiting an exponential rather than a linear growth pattern. Within the 95% CrI, the maximum outbreak size surged from 111 individuals when declared a day after the first death to 523 after a 38-day delay. On the other hand, the number of cases decreases as NPIs level increases, 43 in the minimum ([0,161] 95% CrI) once NPIs level is maximized. When the NPIs level is set as minimum (−50%), the mean number of cases reaches 177 ([0, 585] 95% CrI).

**Fig 7.**
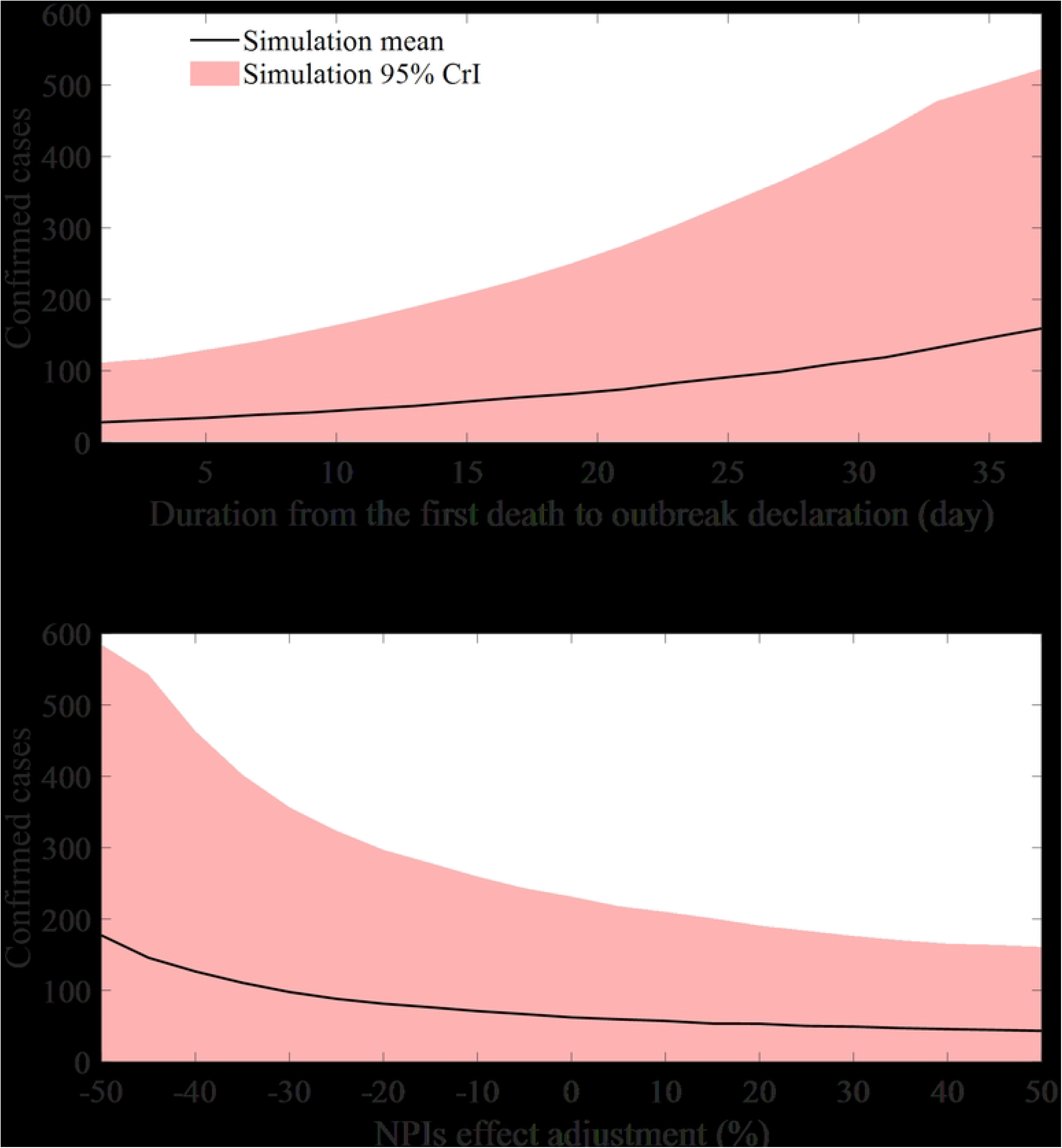
Mean and 95% CrI of confirmed cases considering different factors: Periods leading to outbreak declaration (A), relative intensity of NPIs (B)

Here, we present the outcomes of simulations that concurrently adjust for the previously discussed factors: the timing of outbreak recognition and the intensity of NPIs. Fig 8 maps the NPI intensity on the x-axis against the duration from the first death occurrence to the outbreak declaration on the y-axis. The mean number of cases in each simulation setting is depicted using a color map. For comparison with the baseline scenario outcomes, we integrated contour curves corresponding to the average number of infections in the baseline scenario (yellow, 67) and half (green) and double (red) that count into the graph.

**Fig 8.**
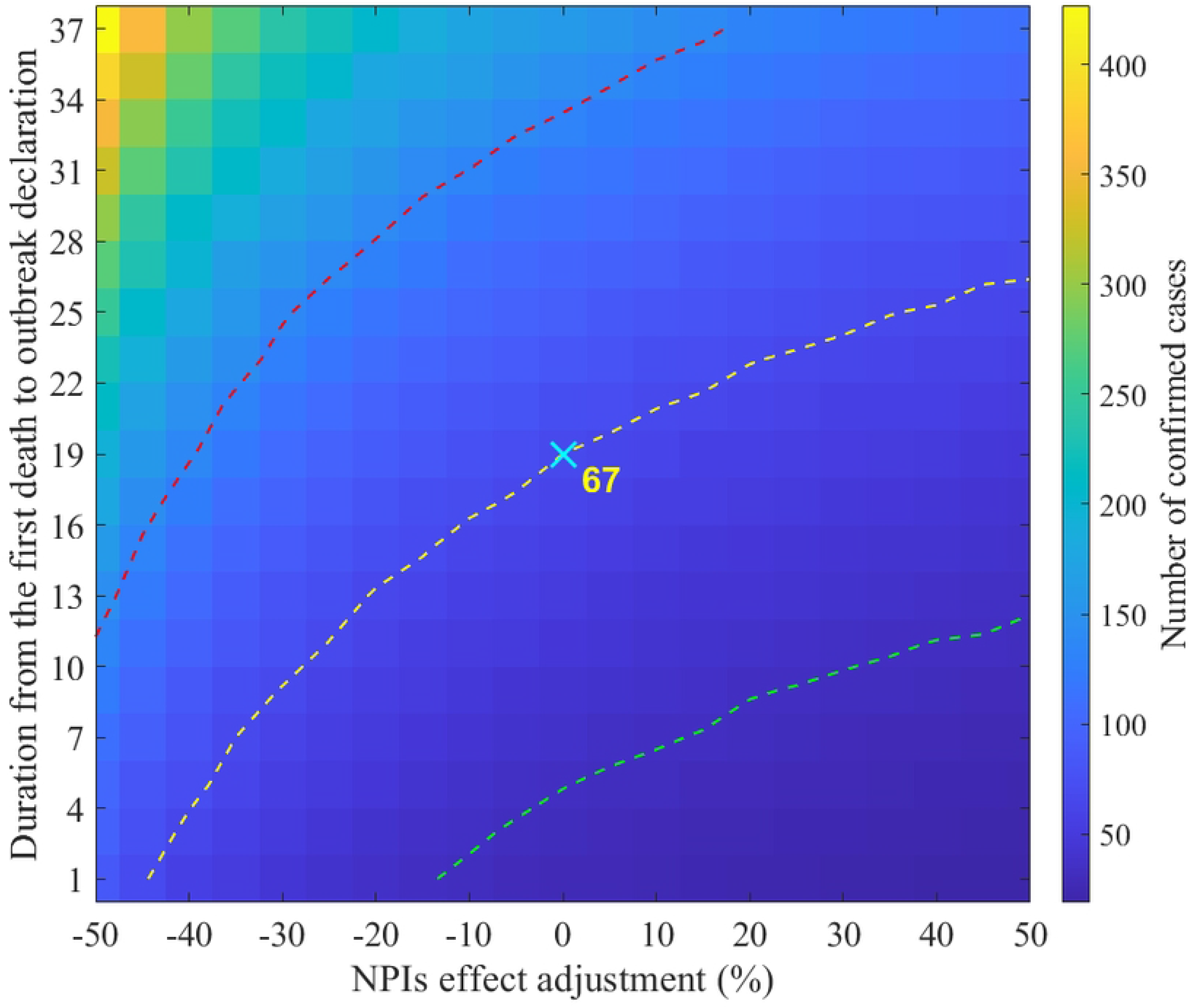
Outbreak scale determined by the intensity of NPIs and the timing of outbreak recognition. Dashed curves represent contours: yellow denotes the outbreak scale from the baseline, whereas green and red indicate half and double the size of the baseline simulation, respectively.

After examining the baseline contour contour, if an outbreak is declared merely a day after the first death, the intensity of the NPIs can be diminished by 45% to attain a similar outbreak magnitude. By contrast, if the outbreak declaration occurs 26 days after the first death, the NPIs must be augmented by 50% to match the baseline outbreak scale. Given a constant NPIs level, outbreak recognition must be advanced by approximately two weeks to cut the infection count by half. Conversely, even with increased NPIs at the baseline recognition juncture, halving the infection scale was impossible.

Let us examine the effect of the vaccination strategy. Fig 9 depicts the mean number of confirmed cases in relation to the timing of when the vaccination is completed. The color of the curves (blue, red, and yellow) represents the proportion of the population that has been vaccinated (10, 20, and 30%). As the vaccination process is expedited or a larger proportion of the population is vaccinated, the number of cases decreases. Conversely, if the vaccination is delayed, the number of cases converges to the number in the baseline scenario (approximately 67). Table 2 lists simulation results. Note that the duration from primary case occurrence to outbreak declaration (*P* 1) was not considered in this table, because NPIs and vaccines are post-outbreak measures.

**Table 2.** Simulation results considering different vaccination rate and duration after outbreak declaration.

| Vaccination rate (%) | Duration (day) | Confirmed cases | | $P2$ duration (day) | |
| --- | --- | --- | --- | --- | --- |
|  |  | Mean | 95% CrI | Mean | 95% CrI |
| 10 | 20 | 42 | [ 1, 157 ] | 99 | [ 15, 194 ] |
|  | 40 | 47 | [ 1, 154 ] | 99 | [ 15, 182 ] |
|  | 60 | 63 | [ 1, 214 ] | 105 | [ 15, 185 ] |
|  | 80 | 67 | [ 1, 236 ] | 108 | [ 15, 191 ] |
| 20 | 20 | 29 | [ 1, 108 ] | 89 | [ 14, 178 ] |
|  | 40 | 40 | [ 1, 123 ] | 94 | [ 14, 170 ] |
|  | 60 | 59 | [ 1, 193 ] | 103 | [ 15, 176 ] |
|  | 80 | 67 | [ 1, 231 ] | 107 | [ 14, 189 ] |
| 30 | 20 | 19 | [ 0, 69 ] | 75 | [ 7, 159 ] |
|  | 40 | 32 | [ 0, 101 ] | 82 | [ 7, 151 ] |
|  | 60 | 53 | [ 0, 190 ] | 93 | [ 7, 165 ] |
|  | 80 | 61 | [ 0, 225 ] | 98 | [ 7, 183 ] |

**Fig 9.**
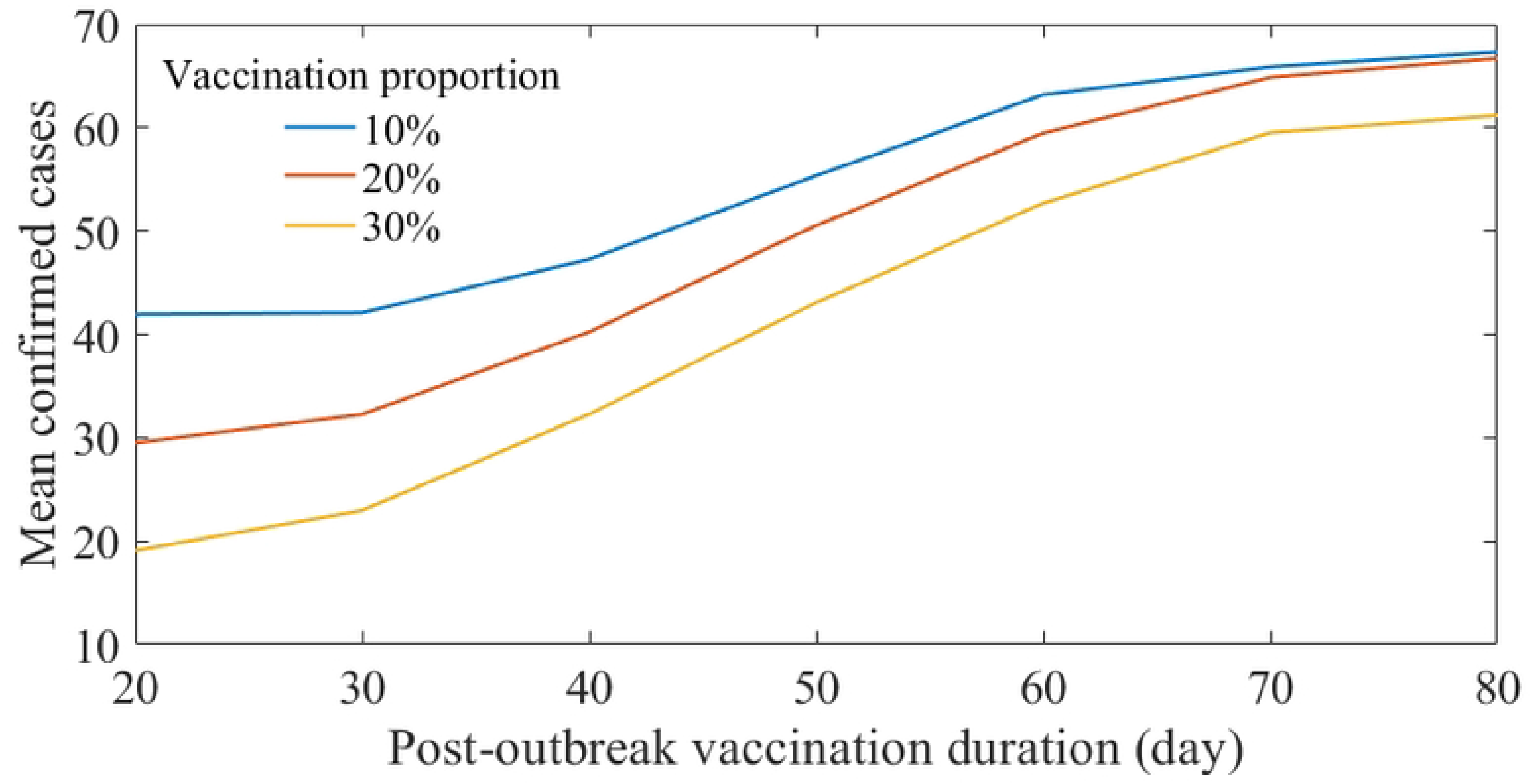
Mean number of confirmed cases considering vaccination strategy. X- and y-axis indicate the duration from outbreak declaration to the vaccination finalizing time and mean number of confirmed cases, respectively.

Similar to what Fig 8 represents, Fig 10 displays mean number of confirmed cases considering vaccination timing and the intensity of NPIs simultaneously. Fig 10A to C contain different simulation results considering various vaccinated proportion of individuals. Fig 10D displays contour curves aggregated from results in Fig 10A to C. Solid (dashed) curves indicate the mean (half mean) number of confirmed cases from the baseline scenario. If the vaccination proportion is set to be 20% and is finalized 50 days after the outbreak declaration, the intensity of NPIs that result in the same number of infections as the baseline scenario was reduced by 40%. Intersection of the blue solid curve and yellow dashed curve indicates the scenario where the confirmed cases could be reduced by half if vaccines can be administered three folds in the same 35 days with 40% eased NPIs.

**Fig 10.**
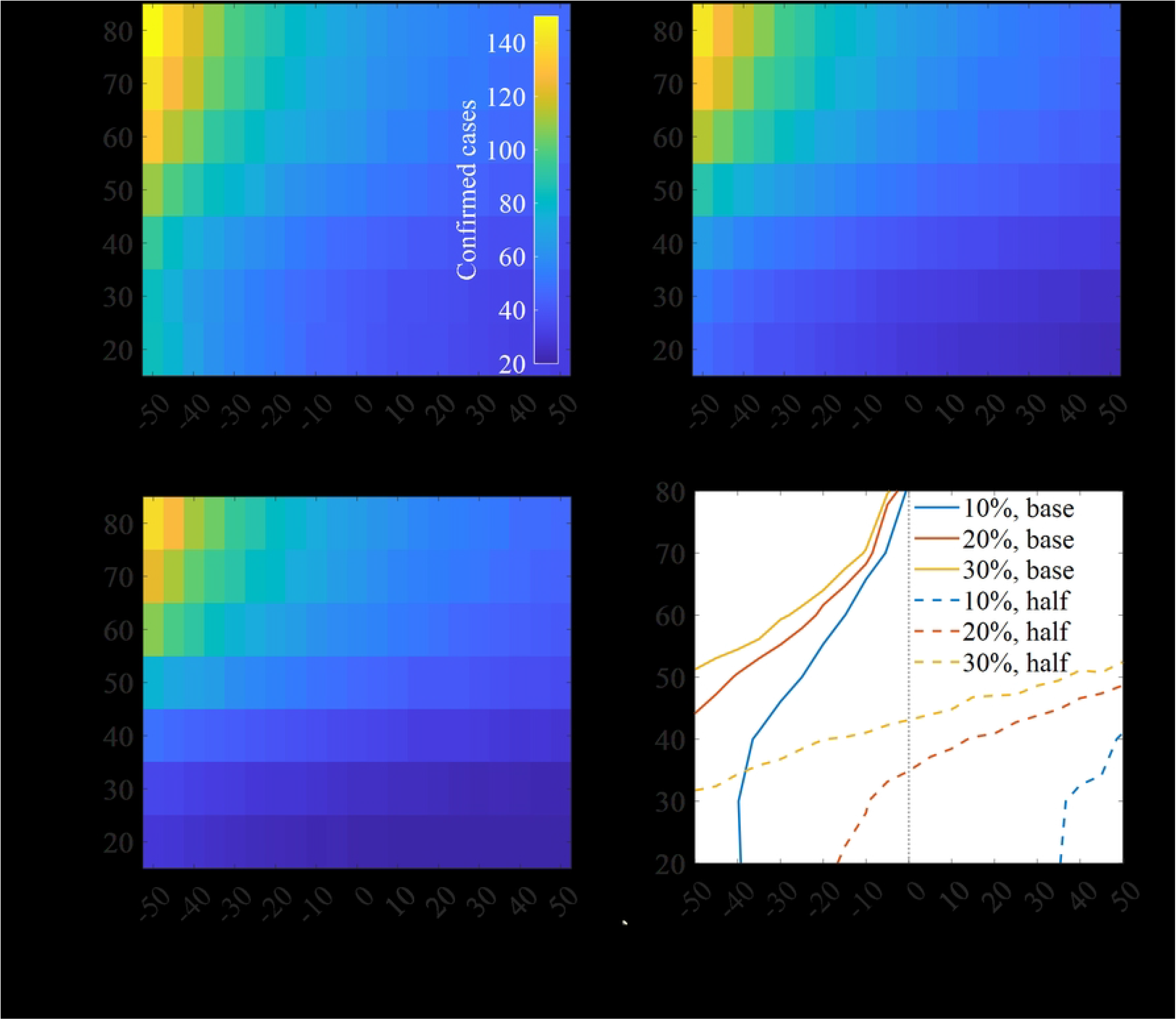
Impact of NPIs and vaccination application. Mean number of confirmed cases where 10%, 20%, 30% of individuals are vaccinated (A-C), contour lines representing the mean number of confirmed cases occurred in the baseline scenario (solid) and half of it (dashed) where the vertical dashed grey line indicates the baseline NPIs intensity (D).

## Discussion

Our model structure, which distinguishes between MS and non-MS as well as reported and unreported cases, provides a detailed understanding of the transmission dynamics. A simple observation of the data reveals a high risk of MS exposure, given the proportion of MS to the total population and the number of infected individuals. The simulation results of our model highlight the high uncertainty of an outbreak, as 95% CrI ranging from 0 to 226 was observed, emphasizing the importance of promptly identifying infected MS once the index case is diagnosed. This underscores the need for enhanced protective measures and training [6]. As Fig 6 indicates, numerous MS could be exposed to the disease, necessitating early and aggressive interventions to identify cases targeting MS.

The simulation of the baseline scenario suggests that the real-world impact of NPIs closely reflects the reduction in the transmission rate and the duration from symptom onset to hospitalization. Furthermore, our simulation results proposed a variety of NPIs that could have been implemented in real-world scenarios. For example, we estimated the effects of NPIs on the transmission rate and infectious period to be 30% and 40%, respectively. However, as Fig 4 demonstrates, these could have been a combination of different values. Our simulation results show the potential to decrease the scale of an outbreak by shortening the infectious period (or reducing the transmission rate), pushing the number towards the upper contours.

The patterns observed in past EVD outbreaks are evident: late detection, inadequate intervention, misinformation, and larger, interconnected populations exacerbate the situation. The West Africa and Kivu epidemics, two significant EVD outbreaks, were the result of these factors. Our model simulation, which did not account for nationwide populations, could not predict an epidemic of that magnitude. However, our simulation still demonstrated exponential growth in the number of confirmed cases as detection was delayed (Fig 7A). On the other hand, in regions with low inter-regional connectivity and population density, small-scale outbreaks could occur even with misdiagnosis/diagnostic delays, as evidenced by the Gabon outbreak in 1994 [43]. In essence, efforts for early detection of EVD spread should not be uniformly distributed across all areas. Instead, if surveillance capacity is strategically focused on areas where the disease is likely to spread, significant effects could be observed.

Our scenario-based study, which varied the timing of the outbreak declaration and the intensity of NPIs (Fig 8), provides valuable insights for policymakers. The results suggest that early recognition and declaration of an outbreak can significantly mitigate the intensity of NPIs required to control the outbreak similarly. In contrast, delays in outbreak recognition necessitate more aggressive NPIs to control outbreak. Localized interventions aimed at identifying confirmed cases among patients with EVD-like symptoms are less burdensome in terms of cost, effort, and manpower requirements than regional lockdowns and nationwide interventions. These findings underscore the importance of early detection.

Vaccine intervention has been observed to significantly reduce the outbreak size, duration, and the burden of NPIs. However, given the storage characteristics of vaccines and the state of medical infrastructure, it is inevitable that the introduction of vaccines will take time. This paradoxically emphasizes the importance of NPIs (Fig 10).

Moreover, even with rapid vaccination, there may be limitations to the vaccine supply. As observed during the Kivu epidemic, when rapid vaccination was implemented, NPIs remained necessary and effective measures. The simulation was conducted based on the Everbo vaccine, which is highly effective with a single dose. However, the Everbo vaccine is effective against the Zaire Ebolavirus, and the case in Uganda involved the Sudan Ebolavirus, not the Zaire strain. This highlights the need for vaccine development, as simulations have shown that the burden of NPIs in future outbreak situations would decrease if a vaccine is available.

This study had several limitations. Firstly, although the MS group was considered separately in the population, the locations where they stay (hospitals or clinics) were not distinguished. Additionally, the vaccination did not reflect the target age of the vaccine. For instance, in the case of Eberbo, the target age was 17 years and older, but this study did not reflect the target age and only used a certain percentage of the total population [44]. Furthermore, the risk of transmission due to the EVD-transmissible semen of recovered patients, found in several cases during past EVD outbreaks, was not reflected [45]. These limitations will be addressed in future work.

## Data Availability

All data produced in the present study are available upon reasonable request to the authors

## Acknowledgments

This research was supported by the Government-wide R&D Fund Project for Infectious Disease Research (GFID), Republic of Korea (grant No. HG23C1629). This paper is supported by the Korea National Research Foundation (NRF) grant funded by the Korean government (MEST) (NRF-2021M3E5E308120711).

